# Cluster Analysis Identified Clinically Relevant Metabolic Syndrome Endophenotypes

**DOI:** 10.1101/2022.11.04.22281926

**Authors:** Aylwin Ming Wee Lim, Evan Unit Lim, Pei-Lung Chen, Cathy Shen Jang Fann

## Abstract

**Aims/hypothesis:** Metabolic syndrome (MetS) is a collection of cardiovascular risk factors; however, the high prevalence and heterogeneity impede proper and effective clinical management of MetS. In order for precision medicine to work for MetS, we aimed to identify clinically relevant MetS sub-phenotypes.

**Methods:** We conducted cluster analysis on individuals from UK Biobank based on MetS criteria to reveal endophenotypes, identified the corresponding cardiometabolic traits and established the association across 21 clinical outcomes. Genome-wide association studies were conducted to identify associated genotypic traits. We further compared the genotypic traits to reveal endophenotypes-specific genotypic traits. Lastly, potential drug targets were identified for the different endophenotypes.

**Results:** Five MetS subgroups were identified which were Cluster 1 (C1): non-descriptive, Cluster 2 (C2): hypertensive, Cluster 3 (C3): obese, Cluster 4 (C4): lipodystrophy-like, and Cluster 5 (C5): hyperglycaemic. Some MetS clusters had higher CVD risks such as C1 (OR=6·765) and C5 (OR=9·486). Despite being non-descriptive across all cardiometabolic traits, C1 had higher risks for most clinical outcomes. MetS clusters also had different risks to various types of cancers. GWAS of each MetS clusters revealed different genotypic traits. *LPCAT2* was associated with all clusters except C2 and expression is specific to immune cells. C1 GWAS revealed novel findings of *TRIM63, MYBPC3, MYLPF*, and *RAPSN*. Intriguingly, C1, C3, and C4 were associated with genes highly expressed in brain tissues: *CN1H2, TMEM151A, MT3*, and *C1QTNF4*. The cluster-specific genotypic traits also revealed potential drug repurposing targets specific to the endophenotypes.

**Conclusion/interpretation:** MetS is highly heterogeneous with endophenotypes that are different in terms of phenotypic and genotypic traits. GWAS of subgroups revealed novel cardiometabolic genotypes which were masked by heterogeneity of MetS.

**Research in context:** *Evidence before this study:* We searched PubMed, Science Direct and Scopus from 1^st^ January 2012 to 30^th^ September 2022 for “unsupervised learning” or “clustering” or “endophenotype” or “subclassifications” or “sub-phenotype” and “metabolic syndrome” or “complex diseases”. Google Scholar, UK Biobank published work and approved research were also searched for similar study. This search only revealed published work in other complex diseases such as T2D (which is heavily referenced in our manuscript), Alzheimer’s diseases, psychiatric diseases, and asthma. None of the previously published work applied the combination of unsupervised learning and GWAS for identification of clinically relevant endophenotypes in metabolic syndrome or any complex diseases.

*Added value of this study:* Metabolic syndrome (MetS) is a known cardiovascular disease risk factor, however the constantly changing MetS criteria and high prevalence of MetS impede proper clinical management of individuals with MetS. Through clustering, we identified MetS endophenotypes with semi-distinctive cardiometabolic traits. Some of the MetS endophenotypes correspond with T2D subgroups discovered by other research groups. However, our endophenotypes are more clinically relevant, due to the differing clinical risks across 21 clinical outcomes. We also identified a non-descriptive MetS subgroup with strikingly high cardiovascular risk which likely to be overlooked in clinical settings. Through genome-wide association studies, our endophenotypes also revealed interesting insights into the genetic causes and biological pathways of MetS. We were able to identified genotypic traits that are unique to each MetS endophenotypes and shared genotypic traits which highlights the common pathophysiology underlying MetS. Lastly, we were also able to reveal potential drug targets for drug repurposing, some drug targets are unique to specific endophenotypes.

*Implications of all the available evidence:* Our study attempted to resolve the issue of MetS heterogeneity, by revealing clinically relevant endophenotypes which might respond to different pharmacotherapy. Furthermore, our findings challenge the “one size fits all” step-wise approach in managing complex diseases, emphasizing tailored treatment for different subgroups of patients, a key step towards precision medicine in clinical practice.

**Graphical Abstract:** 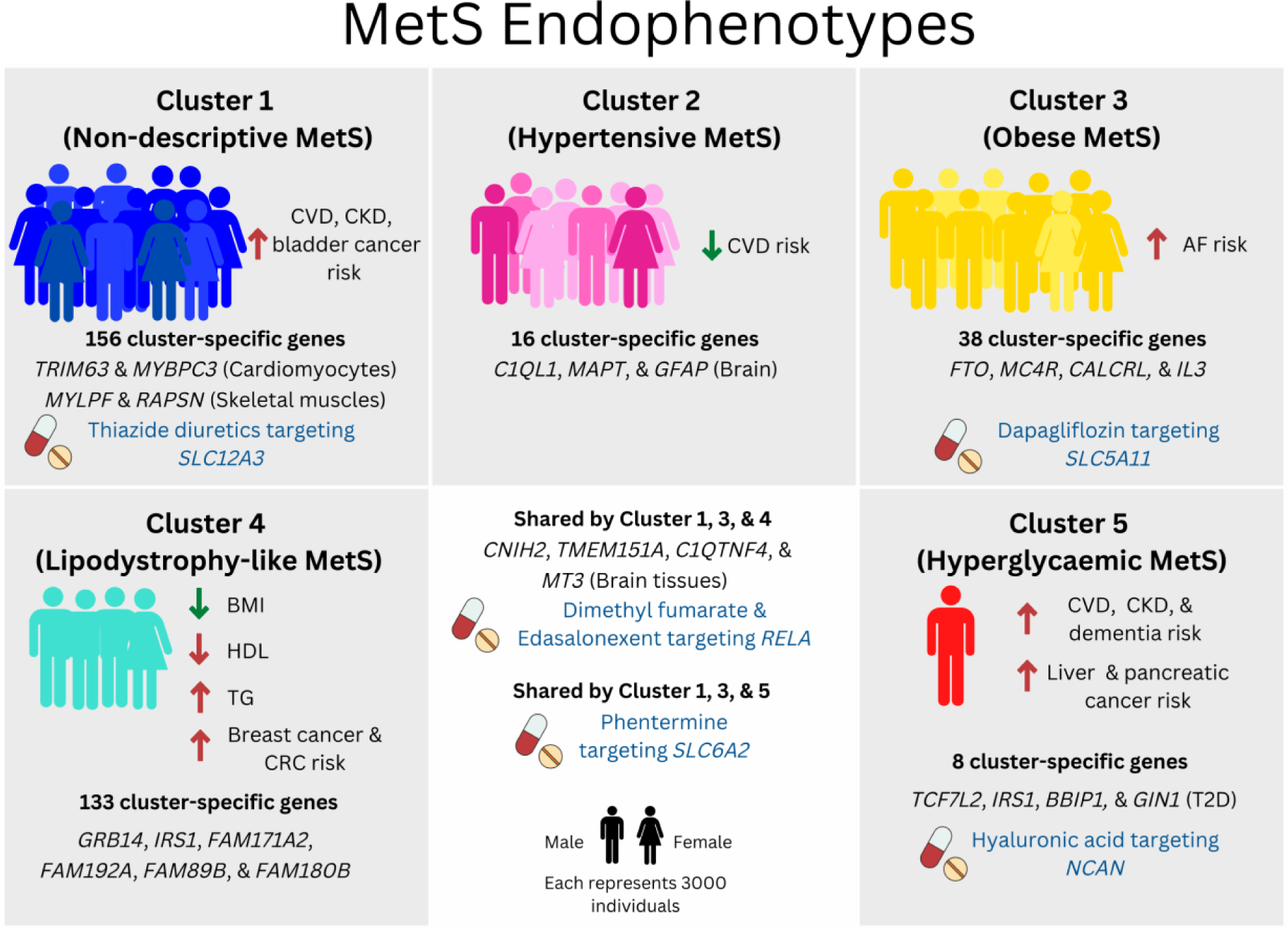

## Introduction

Non-communicable diseases (metabolic diseases, cardiovascular diseases, neurodegenerative diseases, cancer, and pulmonary diseases) are complex; influenced by both genetics and non-genetics factors [1, 2]. To understand the genetic causes and pathophysiology of complex diseases, genome-wide association studies (GWAS) are widely utilized [3-6], however the heterogenous nature of complex disease have impeded effective translation from GWAS to clinical, especially in case-control GWAS of complex diseases [7-9].

Metabolic syndrome (MetS) is the very definition of a highly heterogenous complex disease because MetS encompasses a spectrum of obvolute conditions such as hypertension, dyslipidaemia, type 2 diabetes, and obesity [10, 11]. Diagnosis and classification of MetS is arbitrary and continually evolving according to experts’ consensus and clinical evidence from observational studies [12]. Prevalence of MetS is high and increasing; 34·2% in United States [13], 22·1% in Australia [14] 33·9% in Türkiye [15], and 29·3% in South Korea [16]. With such a high MetS prevalence (almost one third of the general population), managing MetS will expose healthcare system to immense burden.

MetS represents a collection of known cardiovascular risk factors, which contributes to much morbidity and mortality associated with cardiovascular diseases (CVD) such as myocardial infarction, stroke, atherosclerosis, and heart failure [17-19]. In addition to that, MetS causes a multitude of other non-cardiovascular adverse health outcomes: malignancies, renal diseases, and neurological complications (dementia and Alzheimer’s disease) [20-22]. For this reason, identifying individuals with MetS with high risk of complications and preventing MetS-associated diseases is crucial in treating MetS.

However, the binary classification of MetS is insufficient to reflect the heterogeneity, differing risk to disease outcomes and unpredictable pharmacotherapy effectiveness [23, 24]. k-means clustering is a well-known unsupervised learning approach that groups objects with similar characteristics into clusters for identifying subtypes of complex diseases [25, 26]. Ahlqvist *et al* through k-means clustering on type 2 diabetes revealed clinically relevant subtypes of type 2 diabetes in Swedish cohorts [26], which was later replicated in other populations [27-29], albeit with some differences, reflecting population-specific subtypes.

Endophenotype is a concept in genetic epidemiology to describe disease-associated sub-phenotypes with clear genetic connections [30]. For endophenotypes of complex diseases to be clinically relevant, they should be dependent on biological pathways and pathophysiology on top of the ability to distinguish between the heterogeneity of various clinical outcomes. Furthermore, the endophenotypes of complex diseases should allow for stratified or precision treatment approach with differing clinical course and treatment responses. Unsupervised learning should be a useful technique to reclassify MetS, revealing endophenotypes and reducing the heterogeneity of MetS. Furthermore, as MetS is more encompassing which comprises of multiple overlapping cardiometabolic conditions; unsupervised learning when applied on MetS, the bigger picture, might reveal interesting findings.

In our study, we applied unsupervised learning on MetS in UK Biobank (UKB) cohort to reveal clinically relevant endophenotypes of MetS. We further conducted first-of-its-kind GWAS of endophenotypes to identify sub-phenotypes specific associated genotypic traits and potential drug targets.

## Methods

We referred to Strengthening the Reporting of Genetic Association Studies (STREGA) reporting guidelines [31] when drafting the manuscript.

### Study population and phenotypes

UKB is a population-based prospective cohort that recruited 502,637 individuals aged 37–73 from year 2006–2010 across the UK. Full details of the study have been reported in Bycroft *et al* [32]. All participants gave informed consent when joining UKB, which allow for sharing of all anonymized data to authorized researchers. Participants can withdraw consent to sharing of their data at any stages of their participation in UKB. Among the half a million participants, 94.7% individuals are of European ancestry. Based on the ATP III criteria, we classified individuals from UKB as having MetS when fulfilled three or more of the criteria [33, 34]. Individuals were defined as pre-MetS if they met one or two of the MetS criteria.

Based on the Third Report of the National Cholesterol Education Program Expert Panel on Detection, Evaluation, and Treatment of High Blood Cholesterol in Adults (ATP III) criteria [33, 34] :

i. Systolic blood pressure ≥130 mmHg or diastolic blood pressure ≥85 mmHg or on antihypertensive treatment or diagnosed or self-reported to have hypertension
ii. Serum glucose ≥100 mg/dL (5.6 mmol/L) or antidiabetic treatment or diagnosed or self-reported to have T2D
iii. Serum triglycerides ≥150 mg/dL (1.7 mmol/L) or on cholesterol medications
iv. Waist circumference ≥102 cm in men and ≥88 cm in women
v. HDL-C level < 40 mg/dL (1.0 mmol/L) for men and < 50 mg/dL (1.3 mmol/L) for women

Data extraction and transformation was conducted on UKB research analysis platform (RAP) through JupyterLab. Phenotypic data such as anthropometric measurements, cardiometabolic traits, disease status, and mortality are accessed through UKB RAP. Clinical outcomes are defined as shown in Supplementary Table S1 and composite CVD outcome which encompasses CVD defined by ICD10 codes, self-reported conditions, mortality due to CVD, and coronary revascularization procedure as shown in Supplementary Table S2. Missing data was dropped during analysis.

### Unsupervised learning for Mets clusters

K-means clustering is a centroid-based unsupervised learning algorithm that groups a dataset into various clusters by minimizing the within-cluster variances. K-mean clustering algorithm [35] was applied on the standardized MetS criteria of waist circumference, mean arterial pressure (MAP), serum glucose, triglyceride, and HDL cholesterol to identify MetS sub-clusters. The optimal number of clusters, k, was determined through elbow method and silhouette coefficient [36]. For improved initialization of the algorithm, we chose k-means++ over naïve k-means [37]. We also significantly increased the maximum number of iterations to ensure the exploration of entire feature space for elevated accuracy. The above-mentioned was implemented with the class sklearn.cluster.KMeans() from scikit-learn module in Python [38]. One thing to note is that K-means clustering is an NP-hard problem, implementation of other effective metaheuristics needs to be further explored as the sample size increases [39].

### Phenotypic traits comparison among MetS clusters

To compare the phenotypic traits among the different MetS clusters, quantitative traits were compared through one-way analysis of variance and post-hoc analysis with Tukey’s HSD test. The associations of different MetS clusters with clinical outcomes were analysed through age and sex adjusted multivariate logistic regression models; further adjusted for type 2 diabetes status. Statistical significance of the associations with MetS clusters were evaluated using a p-value < 0.001 based on a Bonferroni adjustment for performing 50 tests (21 clinical outcomes and two models: type 2 diabetes-unadjusted and type 2 diabetes-adjusted). We further investigated the role of menopausal status in subset of female individuals from UKB through menopausal status adjusted and unadjusted logistic regression.

### UKB genetic QC and GWAS

Genotyping and imputation of UKB were performed as previously described [32]. GWAS of MetS clusters with healthy control were performed through two-stage REGENIE v3.1.1 [40]. Individuals with ambiguous sex (different sex and genetic sex), sex chromosome aneuploidy, ten or more third-degree relatives identified, non-white British ancestry, who are outliers in heterozygosity and missing rates were removed. Individuals with high genotype missingness (>10%) were also removed. Variants (both genotype and imputation) with high genotype missingness (>10%), low Hardy-Weinberg equilibrium (HWE) p-value (<1×10^−15^), minor allele count (MAC) <20, minor allele frequency (MAF) <0.01, and sample missing rate were filtered out prior to step 1 and 2 of REGENIE [40] using PLINK 2.0 v1.0.6. Imputation information score was set as >0.8.

Step 1 standard logistic regression was conducted with leave-one out cross validation and size of the genotype blocks of 1000 markers. Step 2 was conducted through Firth logistic regression on variants with p-value <0.01 from the standard logistic regression and size of the genotype blocks of 200. Covariates of age, age^2^, first 20 genetic principal components, and sex were included in both steps of REGENIE. Genome-wide significance was determined as p-value <1×10^−9^, multiple testing correction for six GWAS. PLINK and REGENIE are conducted through RAP Swiss Army Knife v4.7.1.

Post-GWAS functional mapping and annotation was conducted through FUMA SNP2GENE and GENE2FUNCTION [41]. Independent significant SNPS were defined with r^2^ threshold of ≥ 0.6 and genome-wide significant p-value of 1 × 10^−9^, a Bonferroni multiple correction for six GWAS from the standard threshold of 5 × 10^−8^; and lead SNPs with a further r^2^ threshold of ≥ 0.1. All candidate SNPs were annotated using built-in ANNOVAR with UKB release 2b 10k White British as reference panel. Annotated SNPs were mapped through positional mapping (physical distances of 10kb, expression quantitative trait locus (eQTL) mapping (SNPs that likely affect expression of genes up to 1Mb), and chromatin interaction mapping with FDR threshold ≤ 1 × 10^−6^. For eQTL mapping, only GTEx v8 tissue types were selected with eQTL maximum p-value ≤ 1 × 10^−3^. Gene mapping are filtered based on functional annotation with CADD score ≥ 12.37, RegulomeDB score ≥ 7, and maximum state of chromatin ≤ 7 from all tissue/cell types available. Tissue-expression analysis of prioritized genes were done through FUMA [41] GENE2FUNCTION using data from GTEx v8 53 specific tissue types. In addition to that, FUMA also performed MAGMA gene and gene-set analysis with gene windows of 10kb. FUMA GENE2FUNCTION also assigned Drug IDs to UnitProt ID of the genes if the genes are targets of the drugs.

### Genotypic traits comparison among MetS clusters

We compared the independent significant SNPs and mapped genes amongst MetS clusters, and defined cluster-specific genotypes as SNPs and genes unique to the cluster. For numerical comparison of the different MetS clusters, we employed Jaccard and cosine similarity coefficient on both independent significant SNPs and mapped genes. We also conducted pairwise genotype comparison (SNPs and genes) of the five MetS clusters with GWAS of all MetS to examine if the GWAS of MetS clusters can reveal novel findings.

## Results

Out of 334,134 individuals of white British ancestry from UKB, 103,996 individuals (31·1%) met at least three of the five MetS criteria and were classified as having MetS. Individuals who met at least one or two of the MetS criteria were classified as pre-MetS (n=184,644, 55·3%).

### Five MetS clusters were identified with distinct phenotypic traits

The optimal number of clusters, k, was determined at five through silhouette coefficient and elbow method (Supplementary Figure S1). Individuals with MetS formed five clusters: Cluster 1 (n=33,707, 32·4%), Cluster 2 (n=23,215, 22·3%), Cluster 3 (n=30,089, 28·9%), Cluster 4 (n=13,116, 12·6%), and Cluster 5 (n=3,869, 3·7%). From the principal component analysis (PCA) plot (Figure 1), the five MetS clusters were located away from the healthy individuals with pre-MetS interspersed among healthy and MetS. MetS Cluster 1, 3, and 4 were in close proximity with some overlaps.

**Figure 1:**
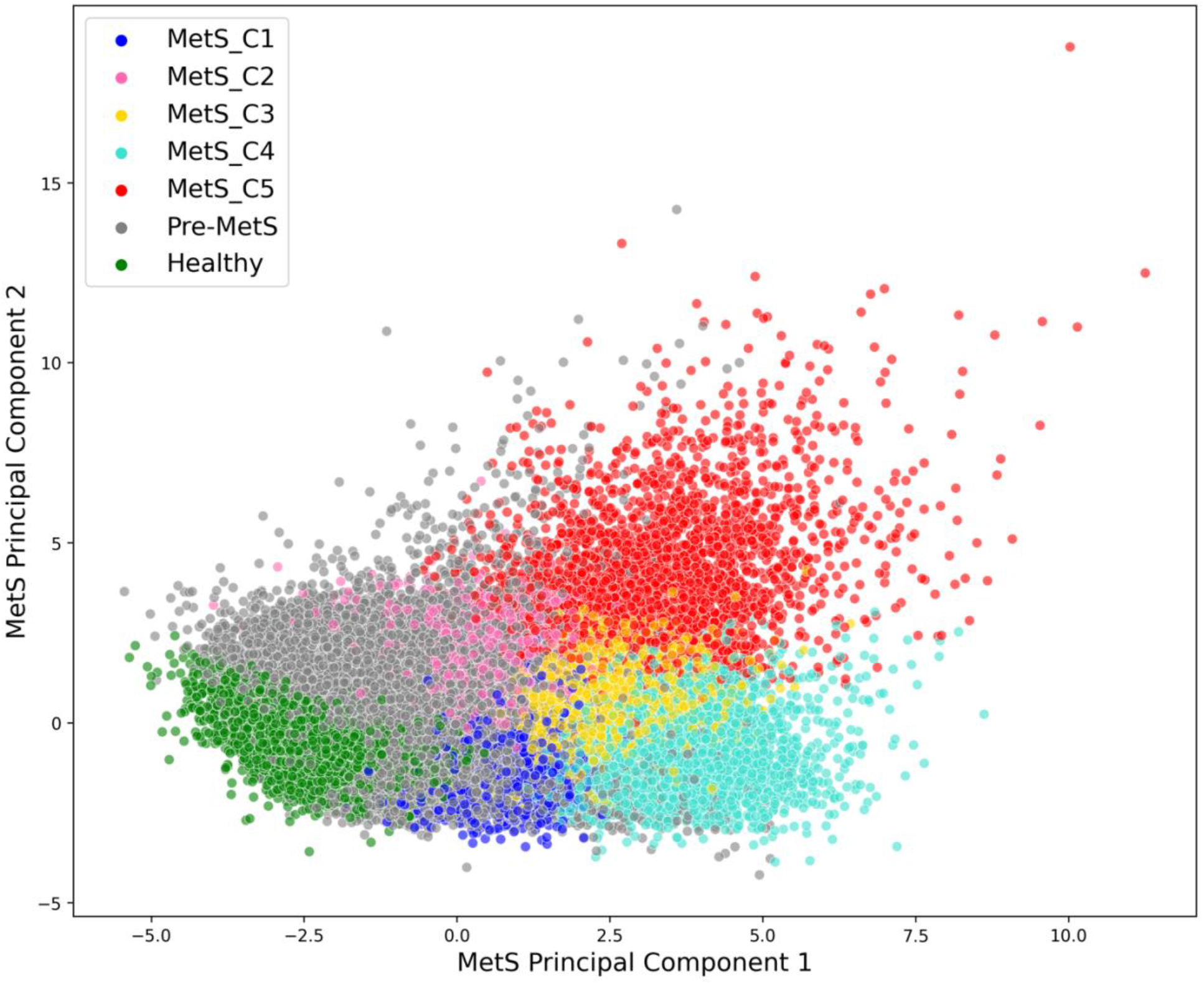
Principal component analysis (PCA) plot of five MetS criteria

MetS clusters were distinctively different in terms of phenotypic traits such as lipid traits, blood pressure, anthropometric measurements, glycaemic traits, and liver enzymes. Cluster 5 (Hyperglycaemic) had the highest glycaemic traits (fasting glucose and HbA_1C_), also reflected in the highest percentage of diagnosed type 2 diabetes status (85·8%) and exogenous insulin supplementation (28·0%). Cluster 4 (Lipodystrophy-like) showed lipodystrophy-like features: highest in triglyceride, lowest in HDL, and more “normal weight” than other MetS clusters as described by Yaghootkar *et al* [42] and Udler *et al* [43]. Cluster 3 (Obese) had the highest obesity traits (waist circumference, hip circumference, BMI, WHR, AVI, and WI). Cluster 2 (Hypertensive) had the highest systolic blood pressure, body fat percentage, and percentage of female (71·5%). Intriguingly, Cluster 2 also had lower lung function as shown by FVC, FEV1, and PEF. Cluster 1 (Non-descriptive) lacked distinctive features when compared to other clusters and generally more similar to the all MetS, pre-MetS, and healthy group. (Figure 1 & 2, Supplementary Table S3) However, it should also be noted that most of the cardiometabolic traits were highly intercorrelated (Supplementary Figure S2), including the phenotypic traits in MetS criteria.

**Figure 2:**
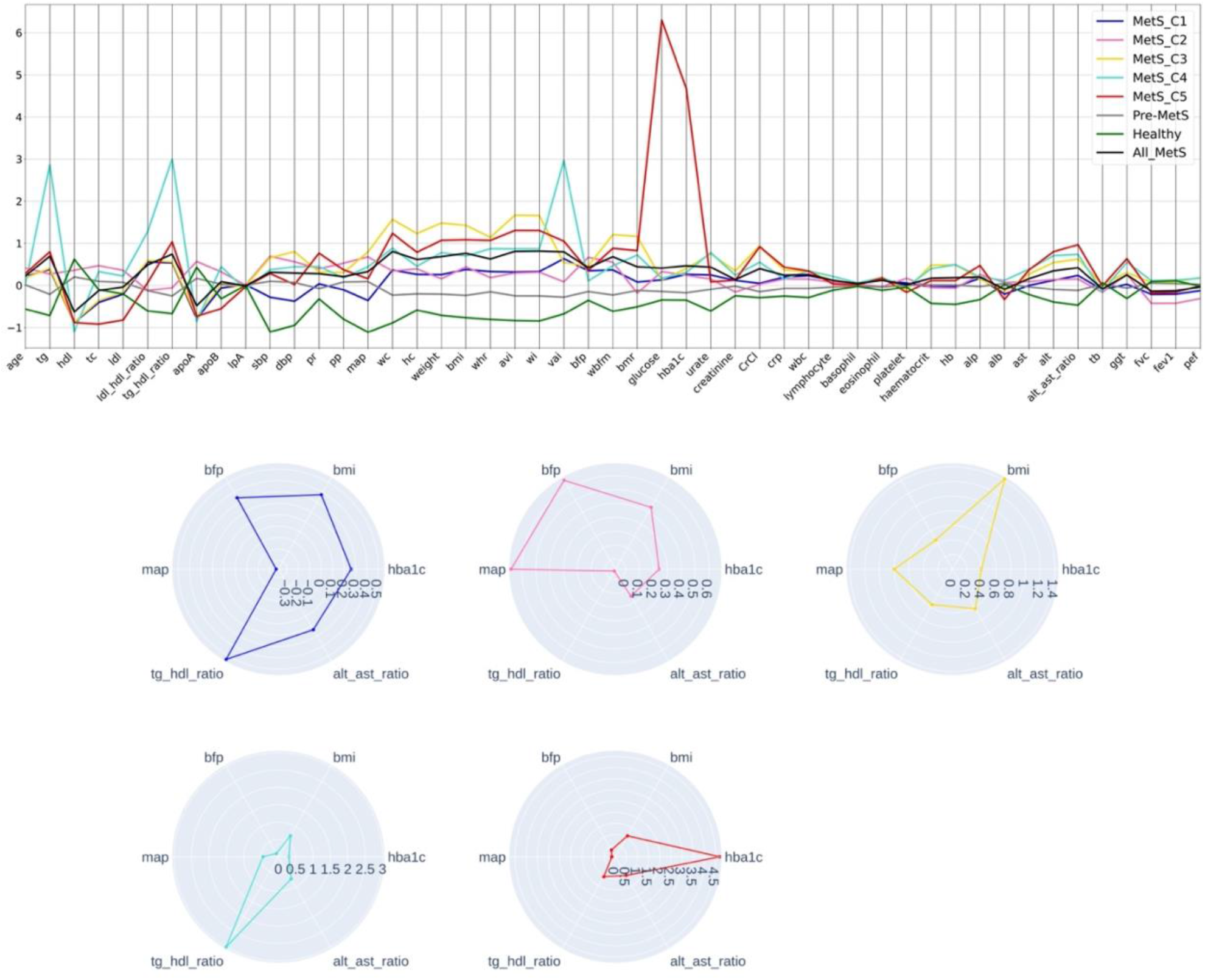
Parallel plot of standard score (Above) for quantitative traits across MetS clusters, pre-MetS and healthy individuals. Radar plot (Below) on features of MetS clusters

### MetS clusters are clinically relevant

Multivariable logistic regression (age and sex adjusted) of each MetS clusters with 21 clinical outcomes such as cardiovascular diseases, stroke, chronic kidney disease, various cancer types, dementia, and Alzheimer’s disease are shown in Figure 3, Supplementary Figure S3, and Supplementary Table S8. MetS clusters had differing clinical risks to various diseases. Overall, MetS is a known CVD risk, our results concurred with a high composite CVD outcomes odds ratio of all MetS, OR=5·517 (5·207, 5·847). Some MetS clusters had higher CVD risks even after adjustment for sex, age, and type 2 diabetes such as Cluster 1 and 5. Despite being undescriptive across the quantitative traits of cardiometabolic risk factors, Cluster 1 had high risks for almost all clinical outcomes even when compared with the all MetS group and the highest risk for composite CVD outcomes after type 2 diabetes-adjustment with OR = 5·633 (5·291, 5·995). Cluster 2 consistently had the lowest risk for all CVD outcomes and with risks similar to that of pre-MetS despite being defined by elevated blood pressure traits. Interestingly, Cluster 3 had the highest risk of AF after adjusting for age, sex, and type 2 diabetes with OR = 2·331 (2·166, 2·507). As expected, the clinical outcome risk of Cluster 5 was mainly due to the hyperglycaemic condition, however, odds ratio remained higher than the collective Mets even after adjusting for sex, age, and type 2 diabetes.

**Figure 3:**
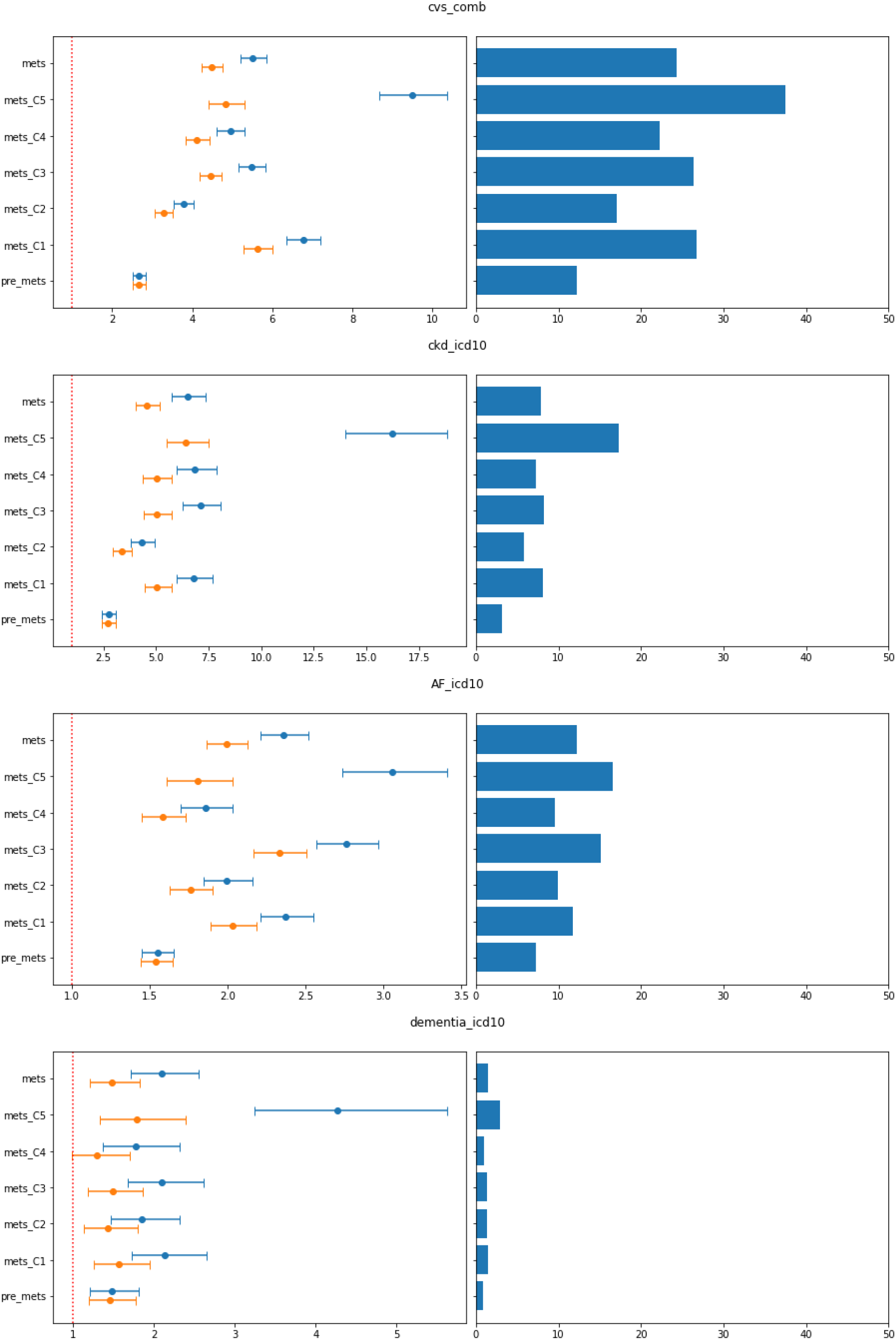
Odds ratio for composite CVD outcomes, chronic kidney diseases, atrial fibrillation and dementia for all MetS, MetS clusters and pre-MetS (Left); blue: adjusted for age and sex; orange: adjusted for age, sex and T2D status. Percentage of cases (Right)

In addition to cardiovascular risks, our results also showed differing elevated risks of various cancer such as liver, breast, pancreas, colorectal, prostate, and bladder cancer among the MetS clusters (Supplementary Figure S3, Supplementary Table S8). MetS is a known risk factor for multiple common cancers [22], however the mechanism linking MetS and cancer is not well elucidated. Most cancer risks for MetS seem to be linked to type 2 diabetes; the association was not significant after type 2 diabetes adjustment, except for breast cancer. Interestingly, the risks for the cancer types investigated differed among the MetS clusters. Cluster 5 had the highest risk for liver cancer OR=5·037 (3·130, 8·101) and pancreas cancer OR=3·487 (2·291, 5·307); Cluster 4 for breast cancer OR=1·413 (1·249, 1·598) and colorectal cancer OR=1·319 (1·140, 1·525); Cluster 1 with highest risk for bladder cancer OR=1·691 (1·392, 2·052).

### Genotypic traits of MetS clusters

GWAS and FUMA SNP2GENE results of each MetS clusters are shown in Figure 4 and more details on independent significant SNPs and prioritized genes in Supplementary Table S11A-F & S12A-F. Each of the GWAS identified multiple candidate SNPs, independent significant SNPs, lead SNPs, and prioritized genes. In terms of mapped genes, the GWAS of MetS clusters also identified cluster-specific genes: 156 out of 481 (Cluster 1), 16 out of 58 (Cluster 2), 98 out of 256 (Cluster 3), 133 out of 461 (Cluster 4), and 8 out of 32 (Cluster 5). Comparison of the Manhattan plots for various GWASs (Figure 5) also revealed some similar and contrasting genomic regions. For example: similar genomic risk loci were identified on chromosome 2 and 16 for all GWAS. Through FUMA GENE2FUNCTION, we can also see that the GWAS of different MetS clusters highlighted genes that are differentially expressed in different tissues (Supplementary Figure S4A-F).

**Figure 4.**
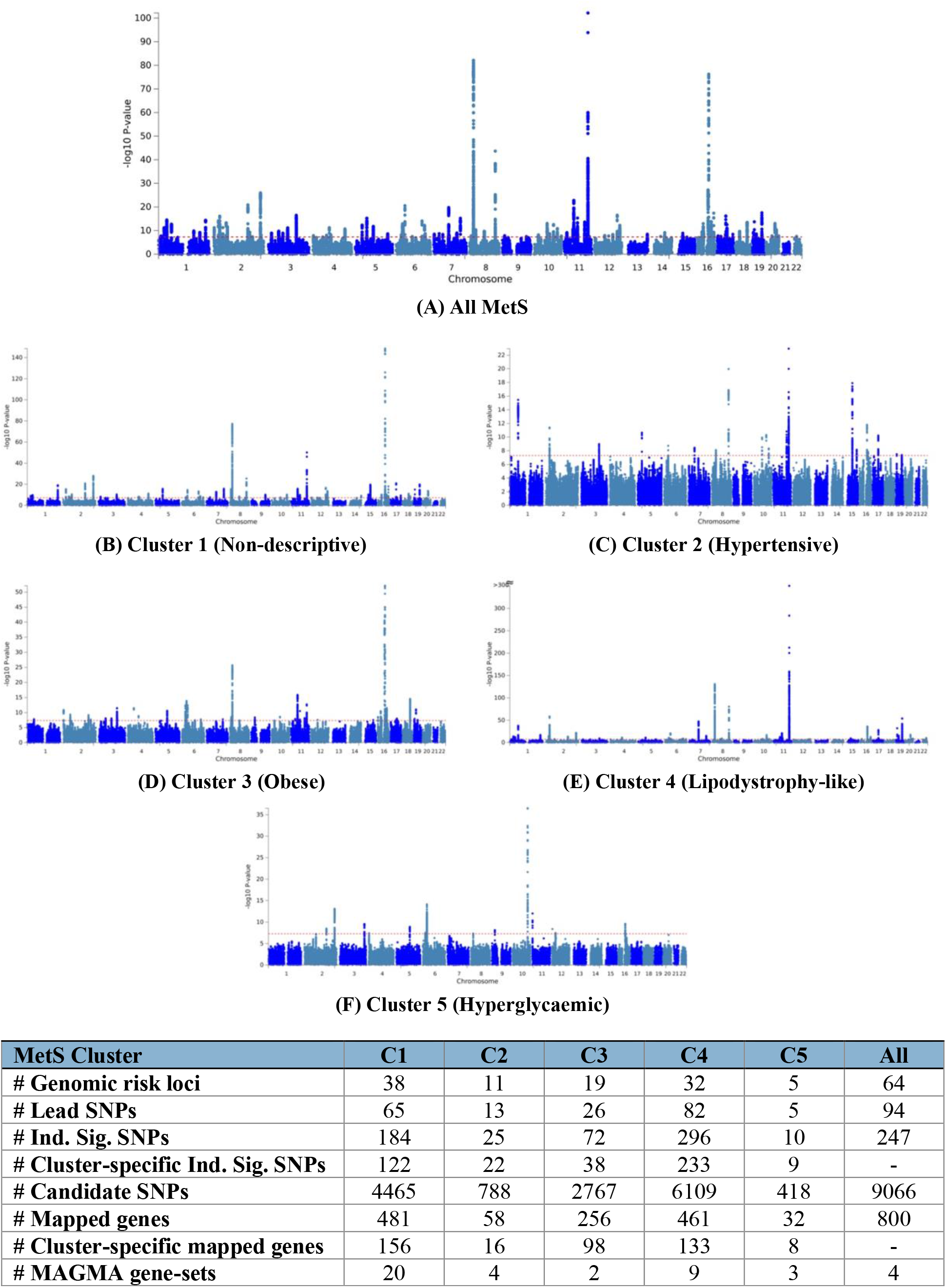
A-F: Manhattan plot for GWAS of all Mets and Mets Cluster 1 – 5

**Figure 5:**
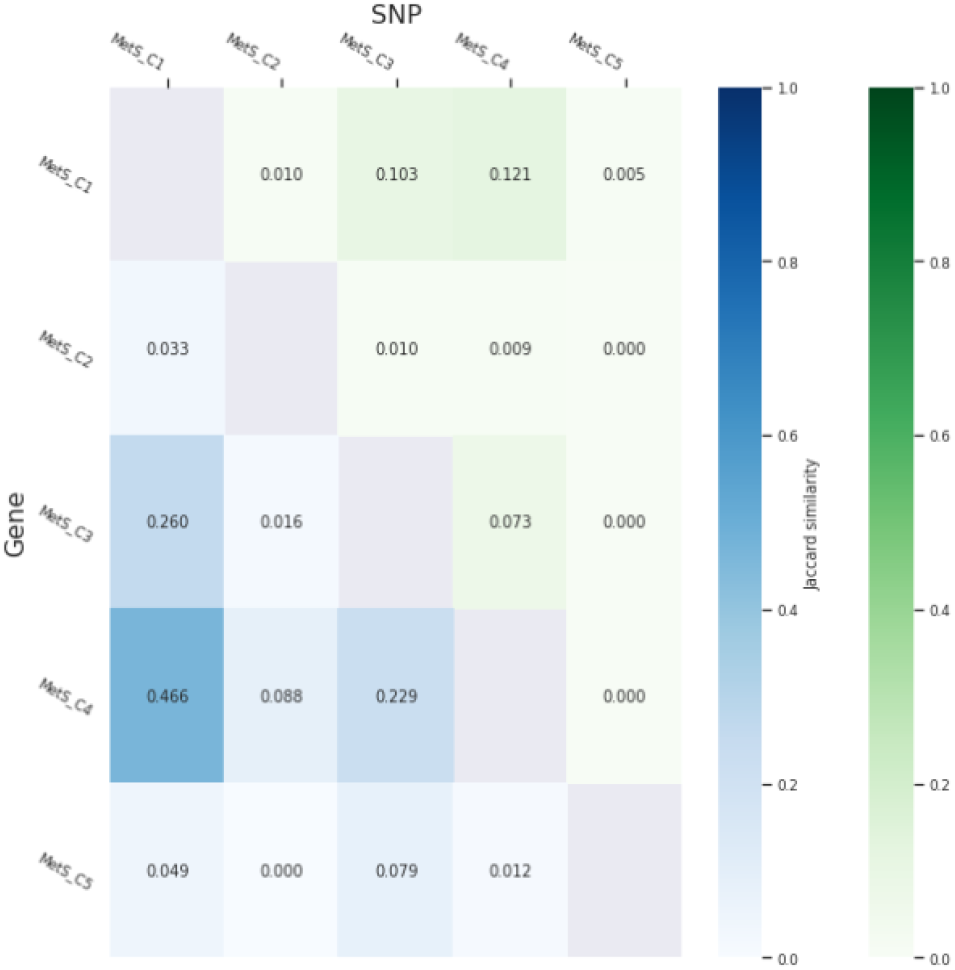
Pairwise genotype comparison of MetS clusters by Jaccard Similarity Index on independent significant SNPs and mapped genes

### Comparison of genotypic traits among clusters

The pairwise genotype comparison among MetS clusters by Jaccard and cosine similarity index on independent significant SNPs and mapped genes are shown in Figure 5 and Supplementary Table S13. MetS Cluster 1, 3, and 4 were the most similar in term of genotypic traits while MetS Cluster 2 and 5 were the most contrasting from other MetS clusters. Furthermore, the comparison of genotypic traits of MetS clusters with all MetS showed that each of the MetS clusters GWAS managed to identified unique SNPs and genes despite them being subgroups of all MetS (Supplementary Table S14).

## Discussion

Through unsupervised learning on MetS criteria, we identified five clinically-relevant MetS endophenotypes which are semi-distinctive in terms of phenotypic and genotypic traits. The clinical relevance of the MetS clusters can be seen across 21 clinical outcomes, allowing for possible risk stratification amongst individuals with MetS. The simple combination of cluster analysis and GWAS allowed important discovery of genotypes underlying the pathophysiology of MetS endophenotypes.

Intriguingly, all MetS clusters except Cluster 2 shared only three genotypes of *LPCAT2* (lysophosphatidylcholine acyltransferase 2), *NUDT21* (nudix hydrolase 21), and *OGFOD1* (2-oxoglutarate and iron dependent oxygenase domain containing 1). *NUDT21* and *OGFOD1* are both been reported to be associated with BMI [44, 45], highlighting the common shared obesity trait that might predispose to MetS in individuals within these clusters. *LPCAT2* has never been reported to be associated with any cardiometabolic traits. Single cell RNA-sequencing data showed high *LPCAT2* expression in immune cells specifically basophil, eosinophil, neutrophil, and monocytes [46, 47]. Furthermore, *LPCAT2* has reported to be responsible for increased expression of inflammatory genes in response to bacterial stimuli [48]. All these previous findings of *LPCAT2* could indicate chronic inflammation being a key player in MetS pathophysiology.

One of the most curious findings is MetS Cluster 1 with no specificity but yet possessed high risks to multiple clinical outcomes including CVD, chronic kidney disease, dementia, and bladder cancer. As a matter of fact, Cluster 1 had the highest CVD risk after adjusting for type 2 diabetes status. The identification of Cluster 1 would be of great clinical importance, as most likely patients who fall within this endophenotype would not be identified as high clinical risk due to the fact that, they are inconspicuous across all cardiometabolic traits. The high clinical risk of Cluster 1 is not merely due to the reported accumulation of risk factors in MetS, as Cluster 1 had higher risk than the all MetS category, but rather some unknown sinister pathophysiology. The prioritized genes of MetS Cluster 1 GWAS could unveil some of the possible underlying mechanisms and biological pathways. MetS Cluster 1 GWAS highlighted *TRIM63* (tripartite motif containing 63), *MYBPC3* (myosin binding protein C3), *MYLPF* (myosin light chain, phosphorylatable, fast skeletal muscle) and *RAPSN* (receptor associated protein of the synapse). *TRIM63* and *MYBPC3* are highly expressed in cardiomyocytes; *MYLPF* and *RAPSN* are highly expressed in skeletal muscles [49, 50]. *TRIM63* was associated with QRS phenotypes, which in turn are linked to cardiac muscle function and hypertrophy, affecting risk of heart failure [51]. *MYBPC3* mutations had been linked to hypertrophic cardiomyopathy in progressive heart failure of both children and adults [52]. Skeletal muscle insulin resistance had been linked to MetS [53] and insulin resistance is a major independent CVD risk factor [54]. The gene expression of skeletal muscle-specific *MYLPF* and *RAPSN* could provide a hint of the possible involvement of insulin resistance in the high CVD risk of Cluster 1.

The phenotypic traits of MetS Cluster 4 are similar to that lipodystrophy-like and liver/lipid type 2 diabetes clusters identified by Udler *et al* [43] with lipodystrophy-like features such as low obesity traits and dyslipidaemia (elevated triglycerides and LDL but reduced HDL). Furthermore, Cluster 4 GWAS highlighted some of the genes previously reported in “lipodystrophy-like” insulin resistance cluster by both Yaghootkar *et al* [42] and Udler *et al* [43] such as *GRB14* (growth factor receptor bound protein 14) and *IRS1* (insulin receptor substrate 1). We also identified several genes from similar gene families such as *FAM* (family with sequence similarity member): *FAM76A, FAM171A2, FAM192A, FAM89B*, and *FAM180B*; similar to *FAM13A* in Yaghootkar *et al* [42]. Cluster 4 supports the importance of identifying individuals of normal weight obesity due to the high CVD risks [55].

MetS and individual MetS components such as hypertension, hyperglycaemia, and obesity are known risk factors for AF [56-60]. Cluster 3 had the highest risk for AF after adjusting for type 2 diabetes status, highlights the alarming risk of developing AF in this MetS endophenotypes, probably due to the role of obesity in MetS towards AF. Cluster 3 GWAS highlighted genes that are uniquely expressed in adipose tissue, brain amygdala, or associated with obesity traits; for example: *FTO* (Fat mass and obesity-associated protein), *MC4R* (melanocortin 4 receptor), *CALCRL* (calcitonin receptor like receptor), and *IL34* (interleukin 34). *FTO* and *MC4R* are well-known obesity genes, which were associated with obesity traits [44, 61] and type 2 diabetes [62, 63]. *CALCRL* have been reported to be associated with obesity traits [64], highly expressed in adipose tissue [49] and negatively associated with leptin, a hormone that help maintain normal body weight [65]. Genetic variants of *IL34* were associated with Alzheimer’s disease [66], BMI [67], and type 2 diabetes in multi-ancestry cohort [68]. However, it is unsure which specific genetic traits underlie the pathophysiology of obesity and AF in this MetS cluster.

We noticed that the intersection of genotypic traits among MetS Cluster 1, 3, and 4 were highly expressed in the brain, specifically hypothalamus and pituitary, such as *CNIH2* (cornichon family AMPA receptor auxiliary protein 2), *TMEM151A* (transmembrane protein 151A), *C1QTNF4* (C1q and TNF related 4), and *MT3* (metallothionein 3). The neuroendocrine systems, especially the hypothalamus and pituitary, are involved in energy thermoregulation and satiety control [69]. Our results highlighted the importance of the neuroendocrine system shared by these three MetS endophenotypes. For instance, *C1QTNF4* modulated food intake patterns and systemic energy metabolism in obese mice [70]. *MT3* expression in hypothalamus of mice may be involved in leptin signalling and peripheral energy expenditure [71].

Despite having higher blood pressure traits, Cluster 2 had lower cardiovascular disease risk, nearer to the risk in the pre-MetS group. We believe that the lower cardiovascular risk in Cluster 2 is due to the higher proportion of female, a cardioprotective factor [72, 73]. Menopausal status especially early menopausal is a cardiovascular disease risk [74]. We noticed that all risks for clinical outcomes were lower when adjusted for menopausal status, indicating that menopausal status seems to be a relevant confounding factor, but the risks still remained significantly higher than the healthy and the pre-MetS group (*Supplementary Table S8 & S9*). However, long-term follow-up might be required to establish the effect of female sex and menopausal status on cardioprotective role in MetS. Furthermore, we lack data on postmenopausal hormone therapy which can affect the cardiovascular risks [75]. Cluster 2 GWAS highlighted some brain-specific genes such as *C1QL1* (complement C1q like 1), *MAPT* (microtubule associated protein tau), and *GFAP* (glial fibrillary acidic protein). *C1QL1, MAPT*, and *GFAP* are known genotypes associated with blood pressure traits [76-78].

MetS Cluster 5 featured the role of type 2 diabetes and hyperglycaemia as a critical component of MetS; also, the strong association with CVD, chronic kidney disease, and neurovascular diseases. Despite being the smallest MetS cluster, GWAS of Cluster 5 managed to identified important well-known type 2 diabetes genes such as *TCF7L2* (transcription factor 7 like 2), *BBIP1* (BBSome interacting protein 1), GIN1 (gypsy retrotransposon integrase 1), and *IRS1* (insulin receptor substrate 1) [62, 79, 80]. The discovery of multiple important type 2 diabetes genes, despite the small sample size of cases and lack of ancestral diversity comparing to previously reported GWASs [62, 79, 80], implied that size does not always matter but the homogeneity of the cases does.

GWAS signals had successfully identify drug targets for various complex diseases such as statins targeting *HMGCR* for lowering low-density lipoprotein [81], ustekinumab and risankizumab repurposing for Crohn’s disease by targeting *IL23R* [82], and antiarrhythmics in AF targeting *SCN5A* [83]. In our study, *SLC12A3* (solute carrier family 12 member 3) was unique to Cluster 1 and is a known drug target for thiazide diuretics [84], which could imply that thiazide diuretics might be a useful drug class for Cluster 1 to reduce CVD risk [85]. *SLC5A11* (solute carrier family 5 member 11), also known as SGLT6, is one of the dapagliflozin target receptors [86]. Inhibition of SGLT6 by dapagliflozin had been reported to reduce oxidative stress, which could be beneficial in reversing diabetic cardiomyopathy [87]. *SLC5A11* was exclusively associated with Cluster 3. *NCAN* (neurocan) which was unique to Cluster 5 had been reported to be associated to type 2 diabetes [68], diastolic blood pressure [88] and cholesterol [89] in previous GWAS. *NCAN* is a drug target of hyaluronic acid, a glycosaminoglycan commonly used in cosmetic treatment, wound healing, and joint pain [90]. Dimethyl fumarate (anti-inflammatory for multiple sclerosis) [91] and edasalonexent (still in clinical trials for type 2 diabetes and Duchenne muscular dystrophy) [92] target *RELA* (proto-oncogene, NF-kB subunit) which was shared by Cluster 1, 3, and 4. Phentermine, widely prescribed to promote weight loss, is an inhibitor of the sodium-dependent noradrenaline transporter encoded by *SLC6A2* (solute carrier family 6 member 2) [93]. *SLC6A2* was specifically associated with Cluster 1, 3, and 5. All these findings might indicate effective drug repurposing for specific MetS endophenotypes.

The major limitation of our study is the Caucasian-ancestry-only analysis which could impact the transferability of endophenotypes identified to other ancestries especially in non-Europeans. However, the lack of heterogeneity in ancestry might have been helpful in our attempt to identify the MetS endophenotypes, which could be masked by mixed ancestries. We expect that the MetS clusters should be applicable for other population as generally subtypes of metabolic diseases based on phenotypic traits are relatively robust across populations [94], the only differences would presumably be the proportion of each endophenotypes. Our assumption would need to be further tested in cohorts of different ancestries. We also expect that there might be population or ancestry specific endophenotypes that are not found in UKB. Furthermore, the hard clustering approach by k-means clustering might not be an ideal approach for identification of diseases subtypes. Nonetheless, overlaps between subgroups in soft clustering might interfere with the assumptions of various statistical tests in subsequent analyses. The clustering based on phenotypic data also possessed an inherent limitation where phenotypic data tends to fluctuate with disease progression and is affected by pharmacotherapy, nevertheless, our MetS clusters still show semi-distinct differences in terms of genotypic data.

## Conclusion

MetS is a highly heterogenous condition with multiple clinically relevant endophenotypes that are semi-distinctive in terms of phenotypic and genotypic traits. The identification of the endophenotypes is a key step towards precision medicine which could allow treatment stratification according to the phenotypic and genotypic traits of endophenotypes.

## Supporting information

Endeavour MetS Supplementary Table S1-S10 S13-S14

Endeavour MetS Supplementary Table S11 & S12

## Data Availability

Analyses in this study was conducted under UKB application ID 46789. The genetic and phenotype datasets are not publicly available but can be accessed via the UK Biobank data access process (http://www.ukbiobank.ac.uk/register-apply/). GWAS summary statistics are available on GWAS Catalog (https://www.ebi.ac.uk/gwas/).

## Contributors

AL, EL, PC, and CF had full access to the study data and take responsibility for the integrity of the data and the accuracy of the data analysis. AL and EL conducted data analysis and was responsible for data visualisation. AL, EL, and CF acquired the funding for the study. PC and CF supervised the project. AL drafted the manuscript, which was reviewed and edited by all authors. All authors gave final approval of the version to be published.

## Acknowledgments

This project is supported by MOST 111-2314-B-001-008 grant from Ministry of Science and Technology of Taiwan. The authors appreciate UKB participants and all staff for their contribution.

## Abbreviations

AF: Atrial fibrillation
ALB: Albumin
ALP: Alkaline phosphatase
ApoA: Apolipoprotein A
ApoB: Apolipoprotein B
AST: Aspartate aminotransferase
AVI: Abdominal volume index
BFP: Body fat percentage
BMI: Body mass index
BMR: Basal metabolic rate
CrCl: Creatine clearance
CRP: C-reactive protein
CVD: Cardiovascular diseases
DBP: Diastolic blood pressure
FEV1: Forced expiratory volume in 1 second
FVC: Forced vital capacity
GGT: Gamma-glutamyl transferase
GWAS: Genome-wide association study
Hb: Haemoglobin
HC: Hip circumference
HDL: High density lipoprotein
LDL: Low density lipoprotein
LpA: Lipoprotein A
MAP: Mean arterial pressure
MetS: Metabolic syndrome
OR: Odds ratio
PEF: Peak expiratory flow
PP: Pulse pressure
PR: Pulse rate
SBP: Systolic blood pressure
SNP: Single nucleotide polymorphisms
TB: Total bilirubin
TC: Total cholesterol
TG: Triglyceride
UKB: UK Biobank
WHR: Waist to hip ratio
WBC: White blood count
WBFM: Whole body fat mass
WI: Waist index

## List of Supplementary Figures

**Supplementary Figure S1:**
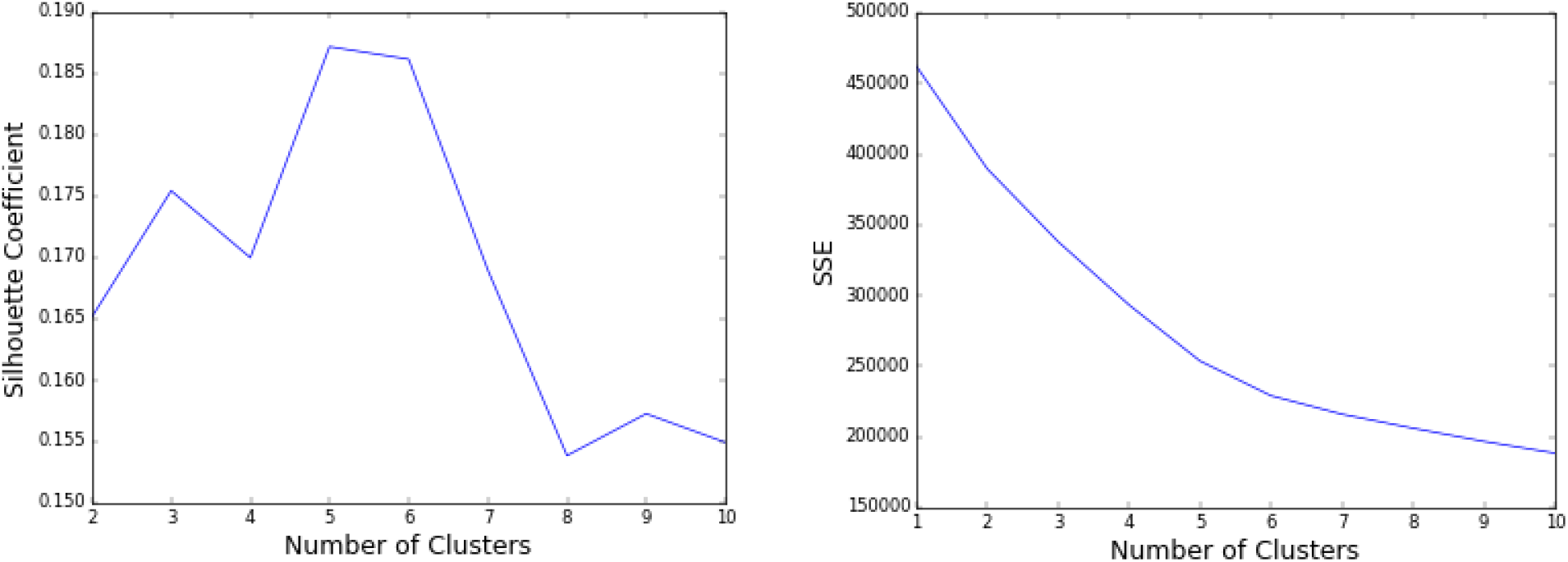
Plot of silhouette coefficient (Left) and within-cluster sum of squared error (SSE) (Right) over number of MetS clusters

**Supplementary Figure S2:**
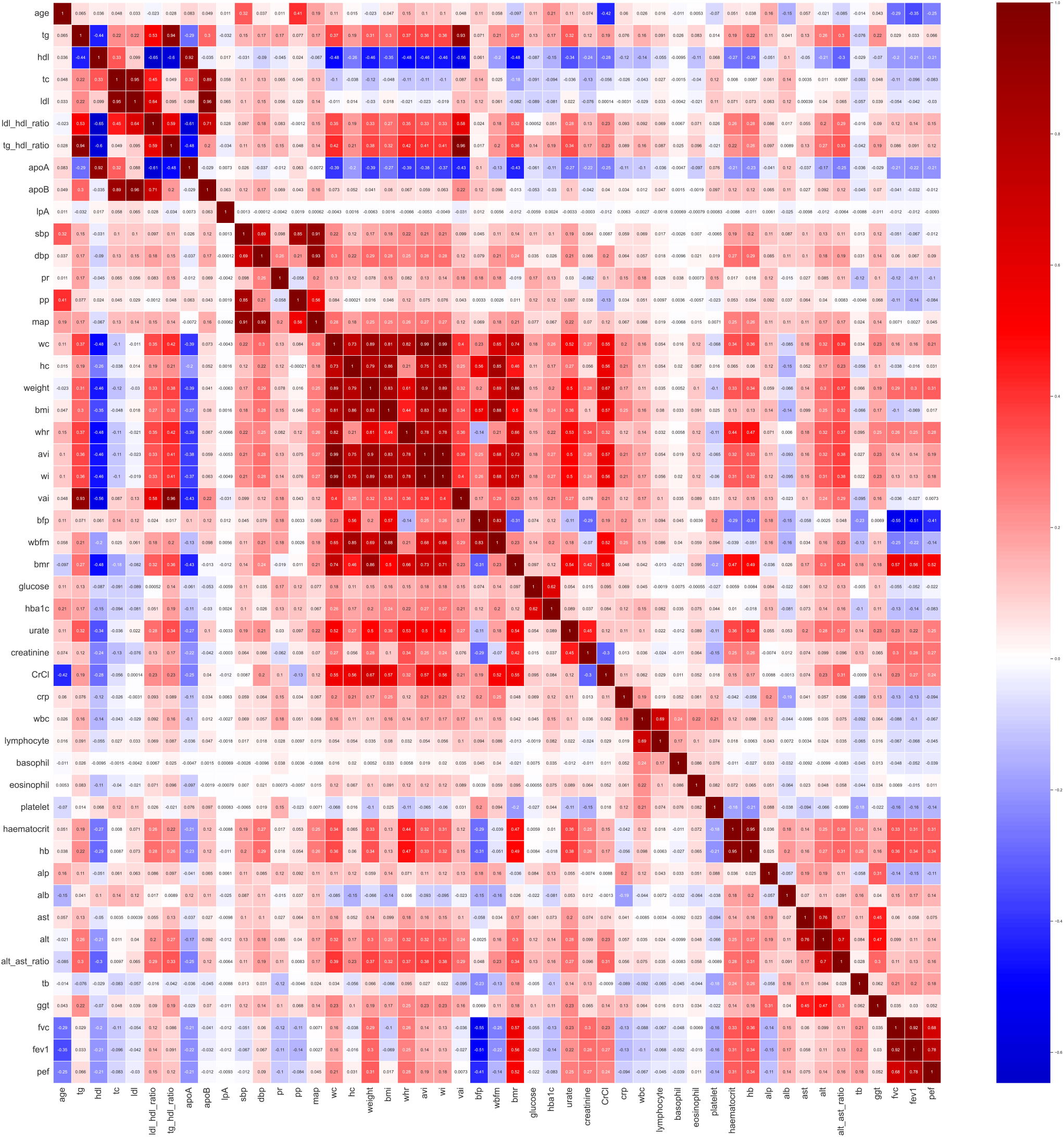
Correlation between phenotypic traits of UK Biobank

**Supplementary Figure S3:**
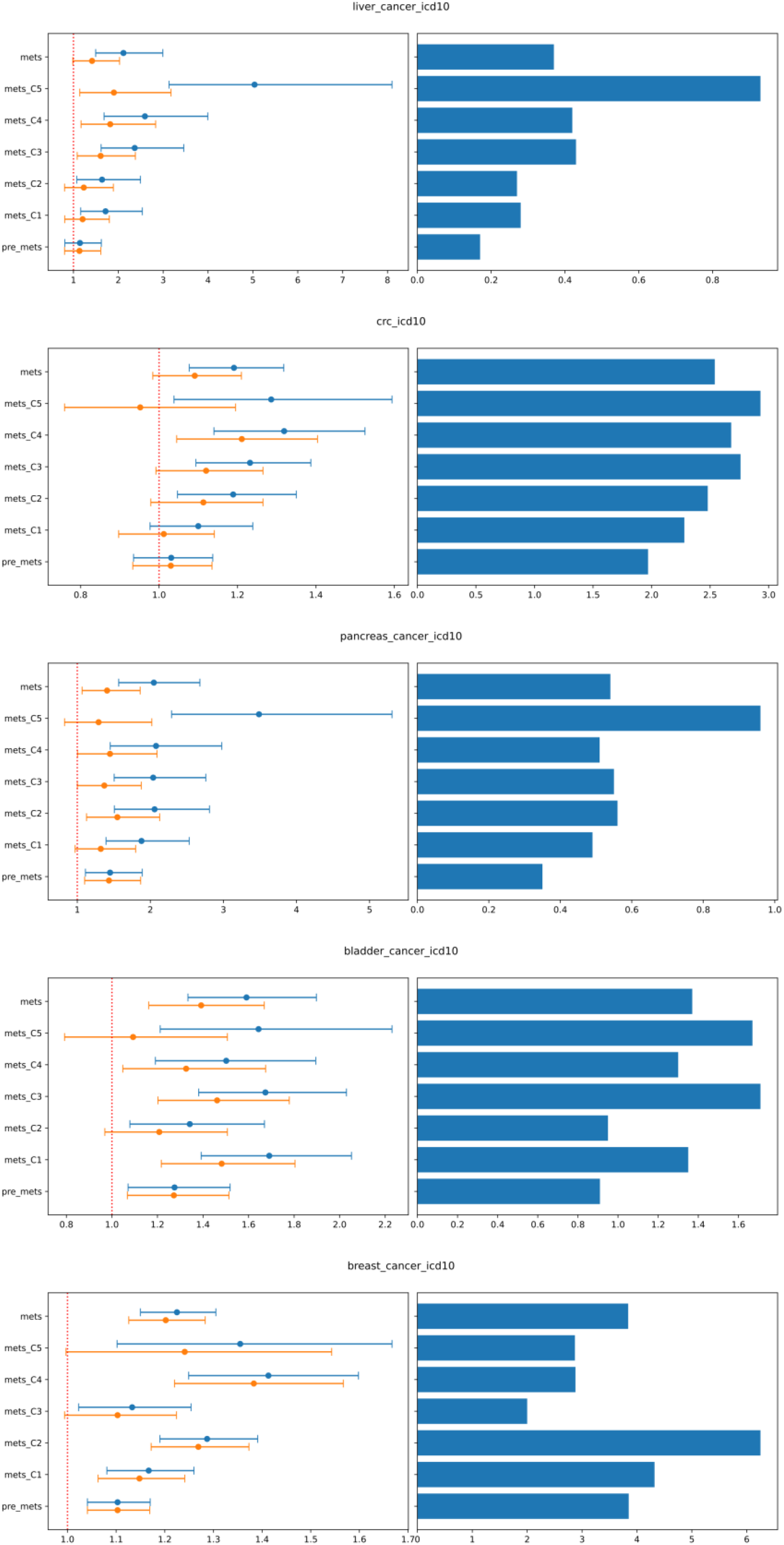
Odds ratio of various cancer types for all MetS, MetS clusters and pre-MetS (Left); blue: adjusted for age and sex; orange: adjusted for age, sex and T2D status. Percentage of cases (Right)

**Supplementary Figure S4A:**
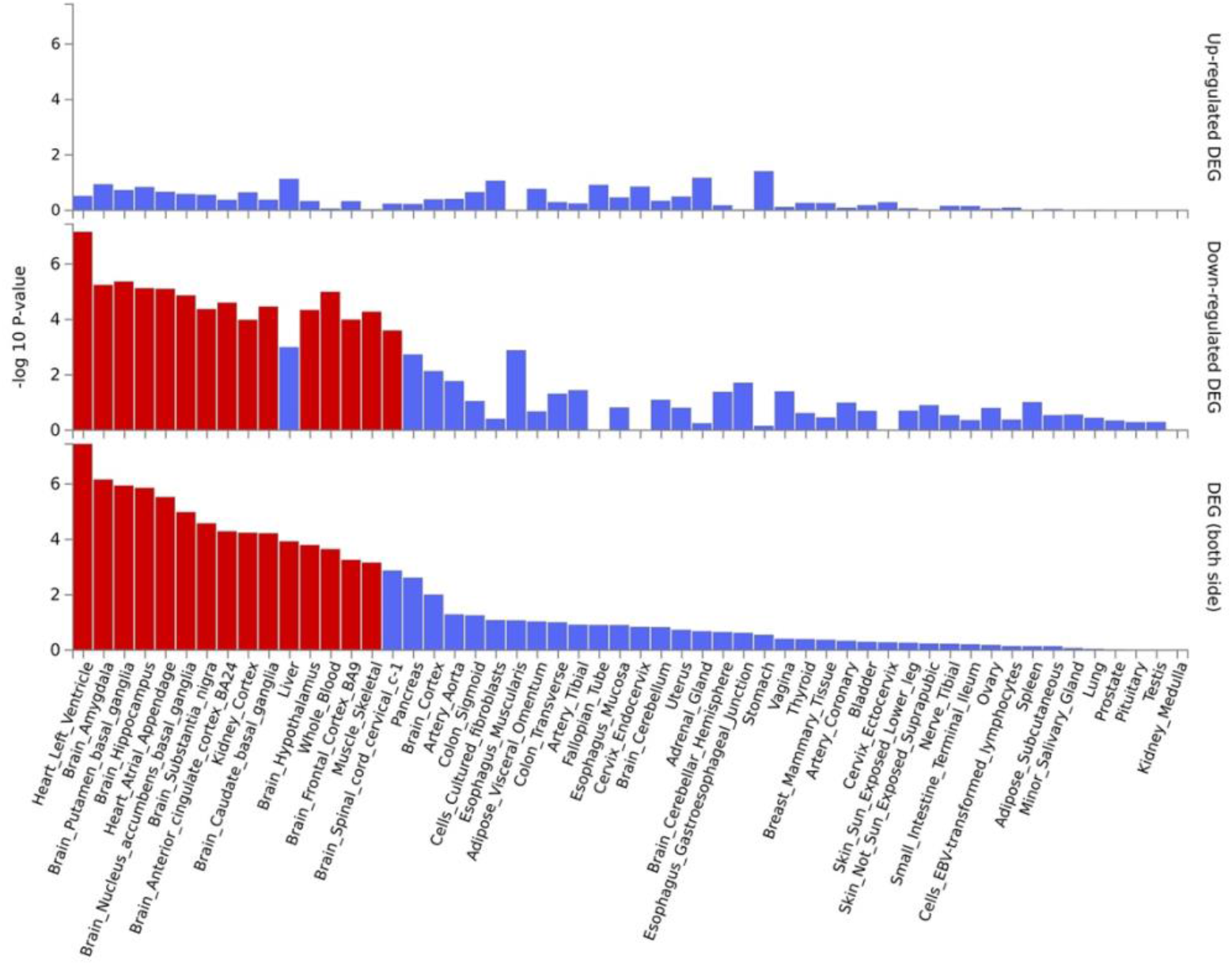
Differentially expressed genes across 53 specific tissue types GTEx v8 for GWAS of all MetS

**Supplementary Figure S4B:**
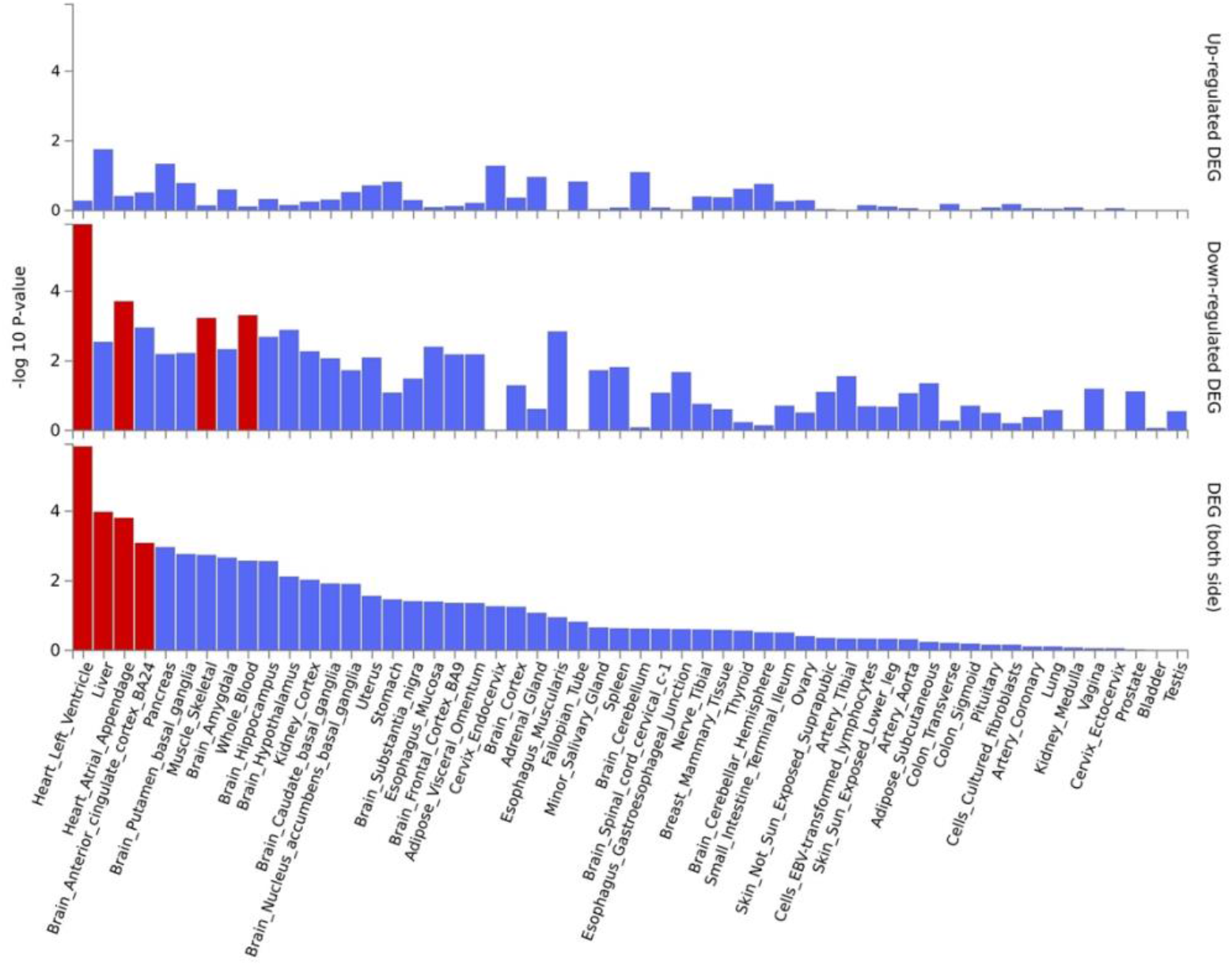
Differentially expressed genes across 53 specific tissue types GTEx v8 for GWAS of Cluster 1 (Non-descriptive)

**Supplementary Figure S4C:**
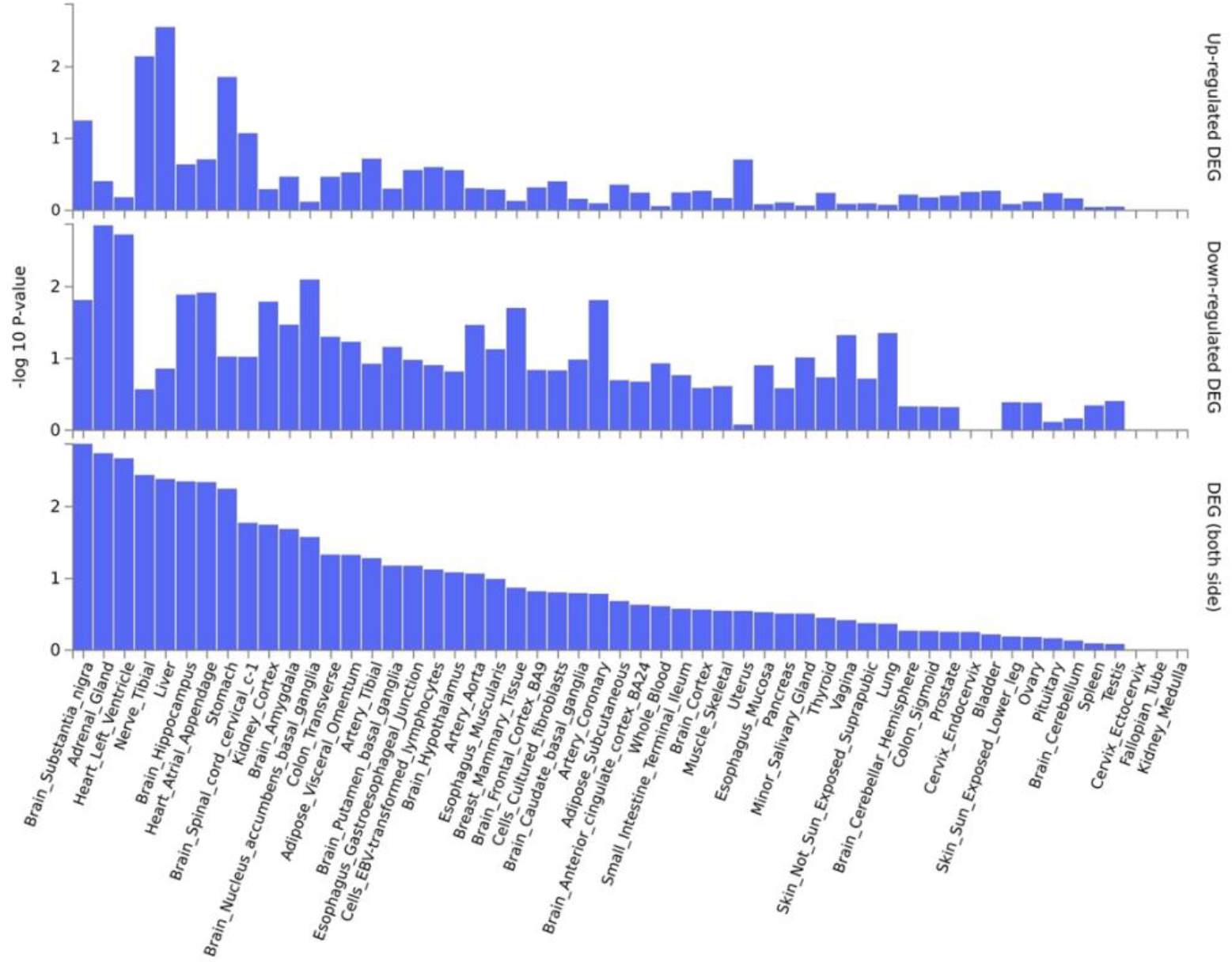
Differentially expressed genes across 53 specific tissue types GTEx v8 for GWAS of Cluster 2 (Hypertensive)

**Supplementary Figure S4D:**
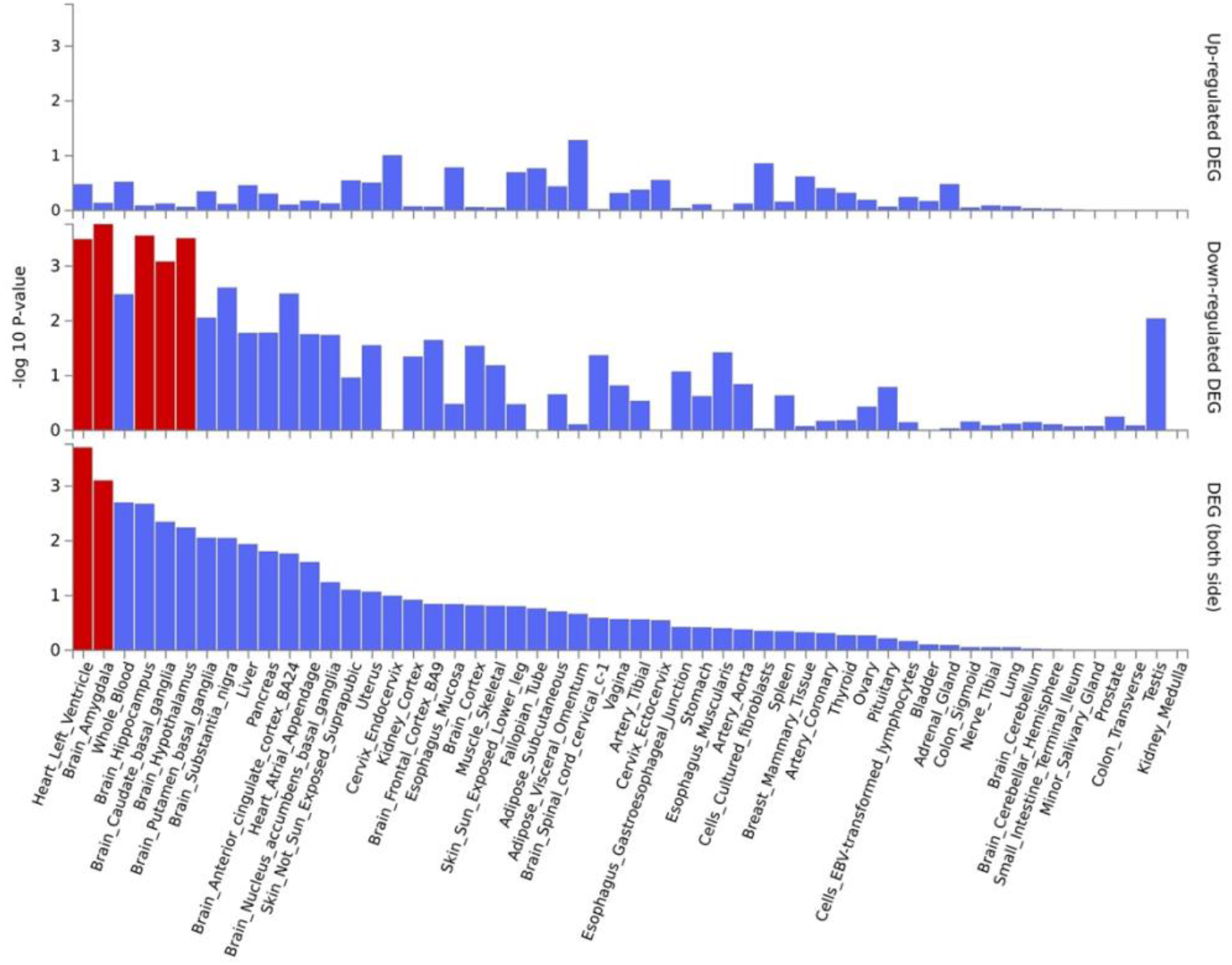
Differentially expressed genes across 53 specific tissue types GTEx v8 for GWAS of Cluster 3 (Obese)

**Supplementary Figure S4E:**
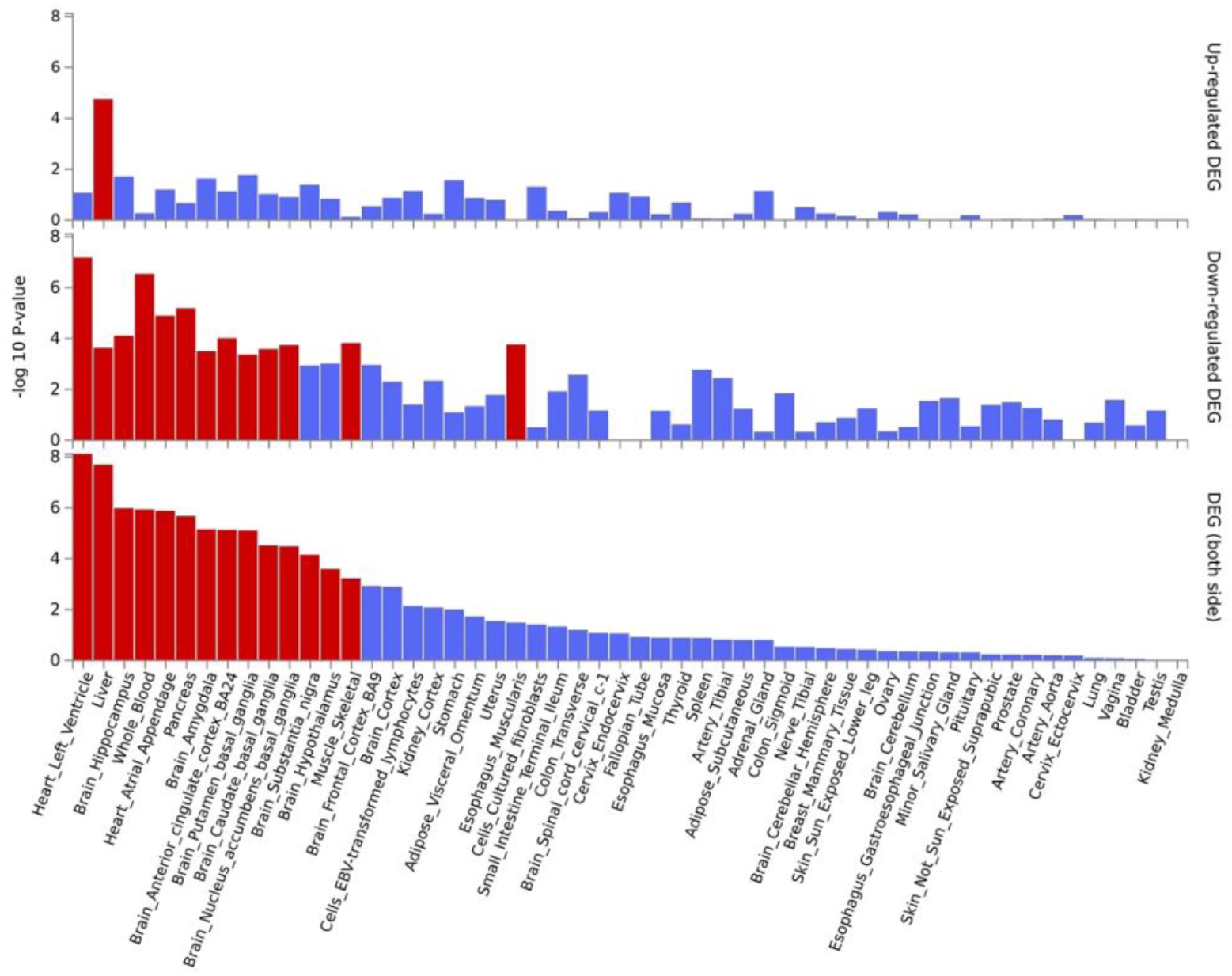
Differentially expressed genes across 53 specific tissue types GTEx v8 for GWAS of Cluster 4 (Lipodystrophy-like)

**Supplementary Figure S4F:**
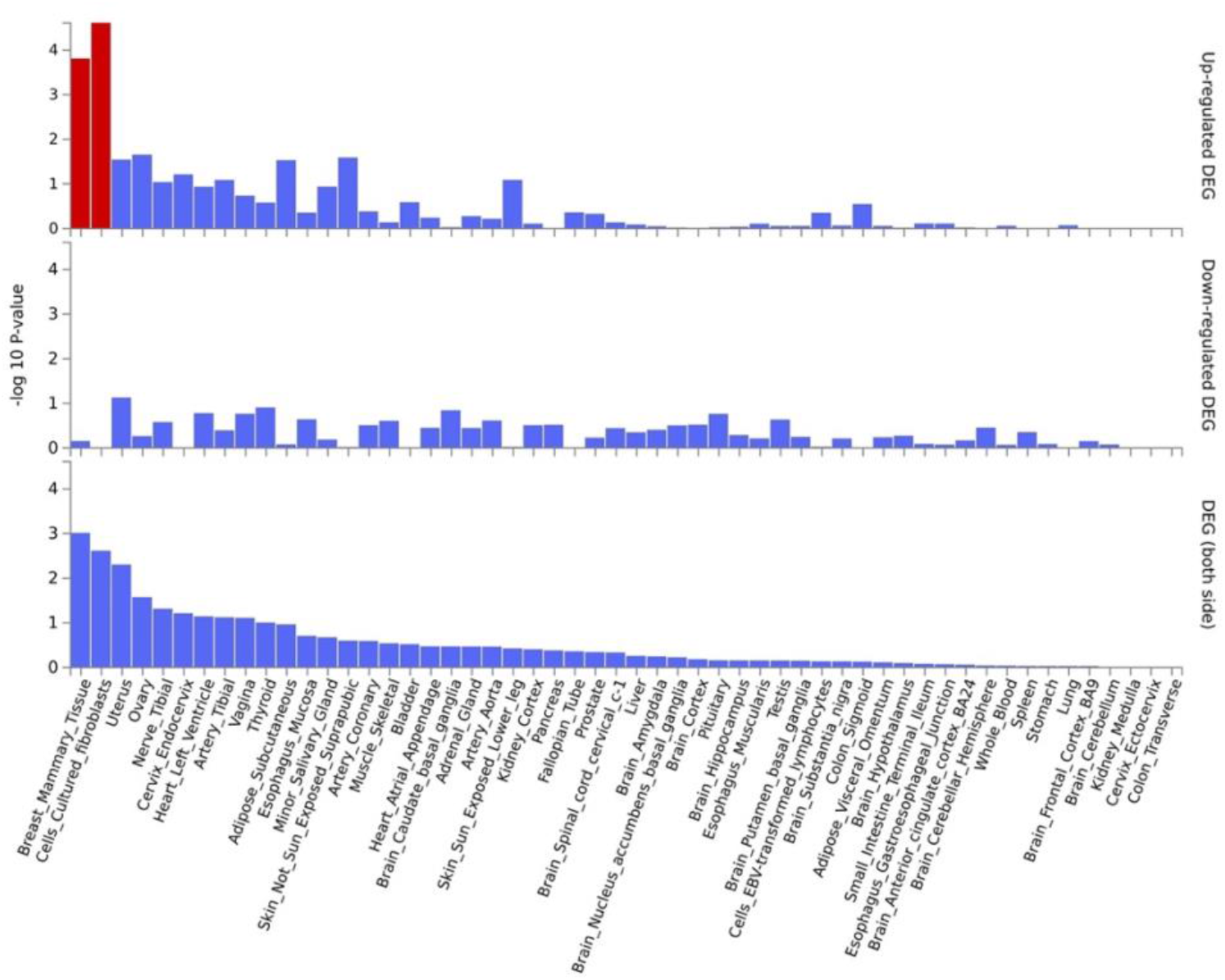
Differentially expressed genes across 53 specific tissue types GTEx v8 for GWAS of Cluster 5 (Hyperglycaemic)

## List of Supplementary Tables

Supplementary Table S1: Definition of clinical outcomes

Supplementary Table S2: Definition of composite cardiovascular outcomes

Supplementary Table S3: Demographics of UKB individuals of European ancestry (Quantitative Traits)

Supplementary Table S4: Standard score of quantitative traits by MetS class

Supplementary Table S5: One-way analysis of variance (ANOVA) of quantitative traits

Supplementary Table S6: Post-hoc analysis of quantitative traits with Tukey’s HSD

Supplementary Table S7: Percentage of categorical traits by MetS class

Supplementary Table S8: Clinical outcomes count, percentage and logistic regression results Supplementary Table S9: Count and percentage of menopausal women

Supplementary Table S10: Clinical outcomes count, percentage and logistic regression results adjusted and unadjusted for menopausal status in women

Supplementary Table S11A-F: Independent significant SNPs from GWAS of MetS clusters Supplementary Table S12A-F: Prioritized genes from GWAS of MetS clusters

Supplementary Table S13: Pairwise genotype comparison of MetS clusters

Supplementary Table S14: Genotype comparison of MetS clusters with all MetS

